# Using Administrative Data to Incorporate Age and Sex-Dependent Resource Use for COVID-19 Acute Care Resource Use Simulations in Ontario, Canada

**DOI:** 10.1101/2020.12.16.20248166

**Authors:** Stephen Mac, Raphael Ximenes, Kali Barrett, Yasin A. Khan, Petros Pechlivanoglou, Juan David Rios, David Naimark, Beate Sander

## Abstract

As the COVID-19 pandemic has progressed, more local data has become available, enabling a more granular modeling approach. In March 2020, we developed a COVID-19 Resource Estimator (CORE) model to estimate the acute care resource use in Ontario, Canada. In this paper, we describe the evolution of CORE2.0 to incorporate age, sex, and time-dependent acute care resource use, length of stay, and mortality to simulate hospital occupancy. Demographics (e.g., age and sex) of infected cases are informed by 4-month averages between March-June, and July-October using 10-year age groups. The probability of hospitalization, ICU admission, and requiring mechanical ventilation are all age and sex-dependent. LOS for each acute care level ranges from 5.7 to 16.15 days in the ward, 6.5 to 10.7 days in the ICU without ventilation, and 14.8 to 21.6 days on the ventilator, depending on month of infection. We calibrated some LOS components to reported ward and ICU occupancy between June 15 and October 31, 2020. Furthermore, we demonstrate the use of CORE2.0 for a regional analysis of Region of Waterloo, Ontario, Canada to simulate the ward bed, ICU bed, and ventilator occupancies for 30 days starting December 2020 for three case trajectory scenarios. Moving forward, this model has become highly flexible and customizable to data updates, and can better inform acute care planning and public measures as the pandemic progresses.

## Introduction

In March 2020, we developed a COVID-19 Resource Estimator (CORE) model to estimate the acute care resource use in Ontario, Canada.(1) This model was based on predominantly international data and made assumptions based on expert opinion due to the limited knowledge of COVID-19 early in the pandemic. Since then, we have updated the model through several iterations (up to CORE 1.8) to better represent Ontario acute care pathways for COVID-19 patients. These updates include updated hospitalization, admissions, length of stay (LOS), and mortality data based on Ontario data, as well as an updated model structure. As the pandemic has progressed, more local, context-specific data on resource use became available, enabling a more granular modeling approach in our latest update to CORE 2.0. The schematic of CORE 2.0 is shown in **Figure 1**. All previous updates can be found in another report.(2) Here we describe the key structural and data updates to CORE 2.0.

**Figure 1.**
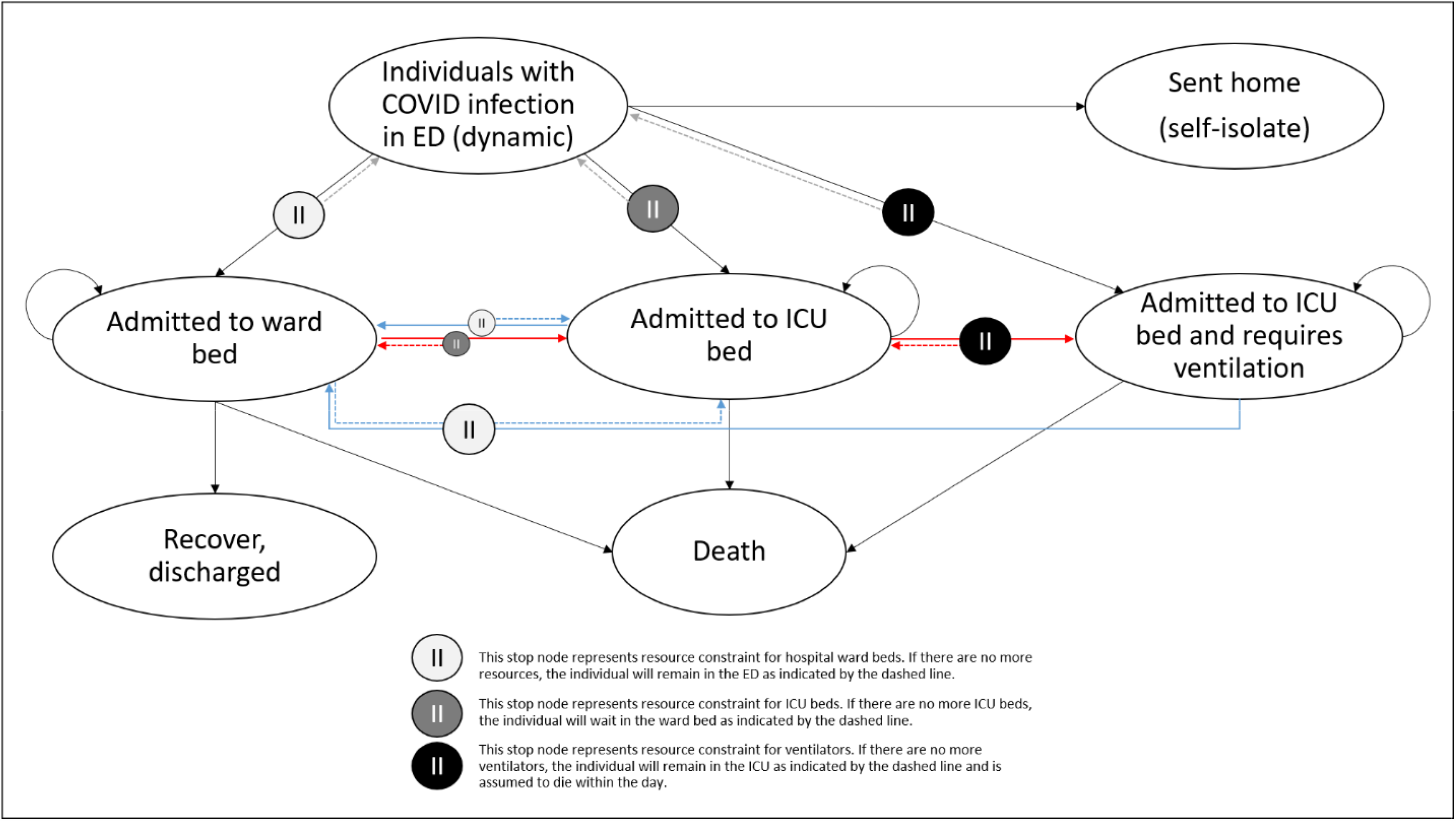
Updated model schematic of CORE 2.0. Blue arrows represent recovery pathways; patients recovering from mechanical ventilation would transition to ward beds or remain in ICU beds (without mechanical ventilator) if there are no ward beds, and patients recovering from the ICU would also recover in the ward post-ICU. Red arrows represent the deterioration of patients who would require more intensive care and treatment. Patients can die in any of the health states.

## Methods

### Evolving COVID-19 Data in Ontario, Canada

CCM Plus is Ontario’s province-wide population-based dataset on all individuals who test positive for COVID-19. (3) CCM Plus includes individual-level data on demographics (e.g., age, sex, region), epidemiology (e.g., likely acquisition), patient characteristics (e.g., co-morbidities), acute care resource utilization (e.g., hospitalization, intensive care unit (ICU) admission, invasive mechanical ventilation (IMV)), health outcomes (e.g., mortality), and long-term care (LTC) residency. We obtained research ethics board approval from the University of Toronto.

We conducted a population-based cohort study using public health data to describe age- and sex-specific acute care use, LOS, and mortality.(4) This study showed that acute care use, including LOS, and mortality varies by age and sex, and has decreased between March and September in Ontario overall and within each groups. We include these changes and temporal trends in the updated model to capture unmeasured effects (e.g., changes in clinical practice).(4)

### Demographics

The modelled population (i.e., incident cases starting March 1, 2020) takes into consideration individuals’ age and sex, informed by Ontario reported COVID-19 case demographics. In CORE 2.0, demographic parameter values (e.g., age and sex) of infected cases are sampled from distributions informed by 4-month averages between March-June, and July-October **(Table 1)**. Day-to-day demographics fluctuate similarly to daily reported cases. We chose 4-months because after June, there was a noticeable increase in the proportion of reported COVID-19 cases in the younger age groups: 0-9, 10-19, 20-29 and 30-39 years. Using this data, an individual’s age and sex now determines the individual’s probability of being hospitalized, requiring ICU admission, mechanical ventilation, and death. A 4-month time frame balanced using specific data and computational resources required for simulation.

**Table 1.** COVID-19 case demographics over time in Ontario.

|  | Proportion (March to June) |  |  | Proportion (July to October) |  |  |
| --- | --- | --- | --- | --- | --- | --- |
| Age (years) | Male | Female | Other | Male | Female | Other |
| < = 9 | 0.0175 | 0.017 | 0.029 | 0.048 | 0.048 | 0.025 |
| 10 to 19 | 0.0325 | 0.034 | 0.122 | 0.108 | 0.11 | 0.1 |
| 20 to 29 | 0.1695 | 0.154 | 0.082 | 0.275 | 0.253 | 0.315 |
| 30 to 39 | 0.1645 | 0.137 | 0.079 | 0.18 | 0.168 | 0.155 |
| 40 to 49 | 0.1525 | 0.147 | 0.167 | 0.128 | 0.135 | 0.088 |
| 50 to 59 | 0.1595 | 0.172 | 0.062 | 0.125 | 0.138 | 0.123 |
| 60 to 69 | 0.1275 | 0.112 | 0.157 | 0.083 | 0.078 | 0.1 |
| 70 to 79 | 0.0795 | 0.069 | 0.059 | 0.033 | 0.033 | 0.03 |
| > = 80 | 0.097 | 0.158 | 0.243 | 0.02 | 0.037 | 0.064 |

Note: Columns sum up to 1; the age breakdown is by sex.

### Time from Episode Date to Hospitalization

In CORE 2.0, we incorporated a lag period of 8 days between symptom onset and hospitalization for all cases. This lag period was informed by the mean number of days from episode date (defined as the earliest of case created, case reported, specimen, and symptom onset dates) to hospital admission date observed in Ontario.(5) We assumed that all COVID-19 individuals do not interact with the healthcare system in this period of 8 days, and do not consider variability across individuals.

### Acute Care Resource Use

The probability of hospitalization, ICU admission, and ICU admissions requiring mechanical ventilation were all updated using age and sex-stratified parameter values **(Table 2)**. For ICU admissions, there was a decrease in the proportion of ICU admissions in September within all age groups, and we therefore used the mean proportion of ICU admissions for September and October for anyone requiring ICU admission post-August.

**Table 2.** Acute care resource use proportions by acute care level and time in Ontario.

|  | **Proportions** |  |  |  |  |  |  |  |  |  |  |  |
| --- | --- | --- | --- | --- | --- | --- | --- | --- | --- | --- | --- | --- |
|  | **Hospitalization, proportion** |  |  | **ICU Admission \|**<br>**Hospitalization**<br>**(March to August)** |  |  | **ICU Admission \|**<br>**Hospitalization**<br>**(September to October)** |  |  | **IMV \| ICU Admission** |  |  |
| **Age**<br>**(years)** | **Male** | **Female** | **Other** | **Male** | **Female** | **Other** | **Male** | **Female** | **Other** | **Male** | **Female** | **Other** |
| < = 9 | 0.02 | 0.01 | 0 | 0.17 | 0 | 0 | 0.50 | 0.14 | 0 | 0.50 | 0 | 0 |
| 10 to 19 | 0.01 | 0.01 | 0 | 0.24 | 0.06 | 0 | 0.20 | 0.25 | 0 | 0.20 | 0 | 0 |
| 20 to 29 | 0.01 | 0.02 | 0.02 | 0.15 | 0.18 | 1.00 | 0.06 | 0 | 0 | 0.73 | 0.56 | 1.00 |
| 30 to 39 | 0.03 | 0.03 | 0 | 0.20 | 0.14 | 0 | 0.18 | 0.15 | 0 | 0.59 | 0.41 | 0 |
| 40 to 49 | 0.06 | 0.05 | 0.03 | 0.29 | 0.19 | 1.00 | 0.17 | 0.10 | 0 | 0.59 | 0.62 | 1.00 |
| 50 to 59 | 0.13 | 0.07 | 0.05 | 0.38 | 0.31 | 0 | 0.24 | 0.17 | 0 | 0.64 | 0.63 | 0 |
| 60 to 69 | 0.21 | 0.15 | 0.06 | 0.37 | 0.26 | 0 | 0.33 | 0.31 | 0 | 0.61 | 0.70 | 0 |
| 70 to 79 | 0.35 | 0.27 | 0.05 | 0.29 | 0.20 | 0 | 0.30 | 0.12 | 0 | 0.63 | 0.59 | 0 |
| > = 80 | 0.34 | 0.21 | 0.04 | 0.09 | 0.07 | 0.25 | 0.13 | 0.10 | 0 | 0.43 | 0.50 | 0 |

ICU, intensive care unit; IMV, intensive mechanical ventilator

### Mortality

The probability of death for patients in each of the acute care levels were updated by age, sex, and time **(Table 3)**. Similar to the case demographics, there was a notable change in mortality, overall, and within age groups, in May and June. Hence, we used the mean mortality from March to May for the first three months of the pandemic, and the mean mortality from June to October for incident cases from June onwards. To parameterize the model, which is in daily time-steps, mortality was converted over the duration of the average LOS in each respective level of acute care (ward, ICU without IMV, and ICU with IMV).

**Table 3.**
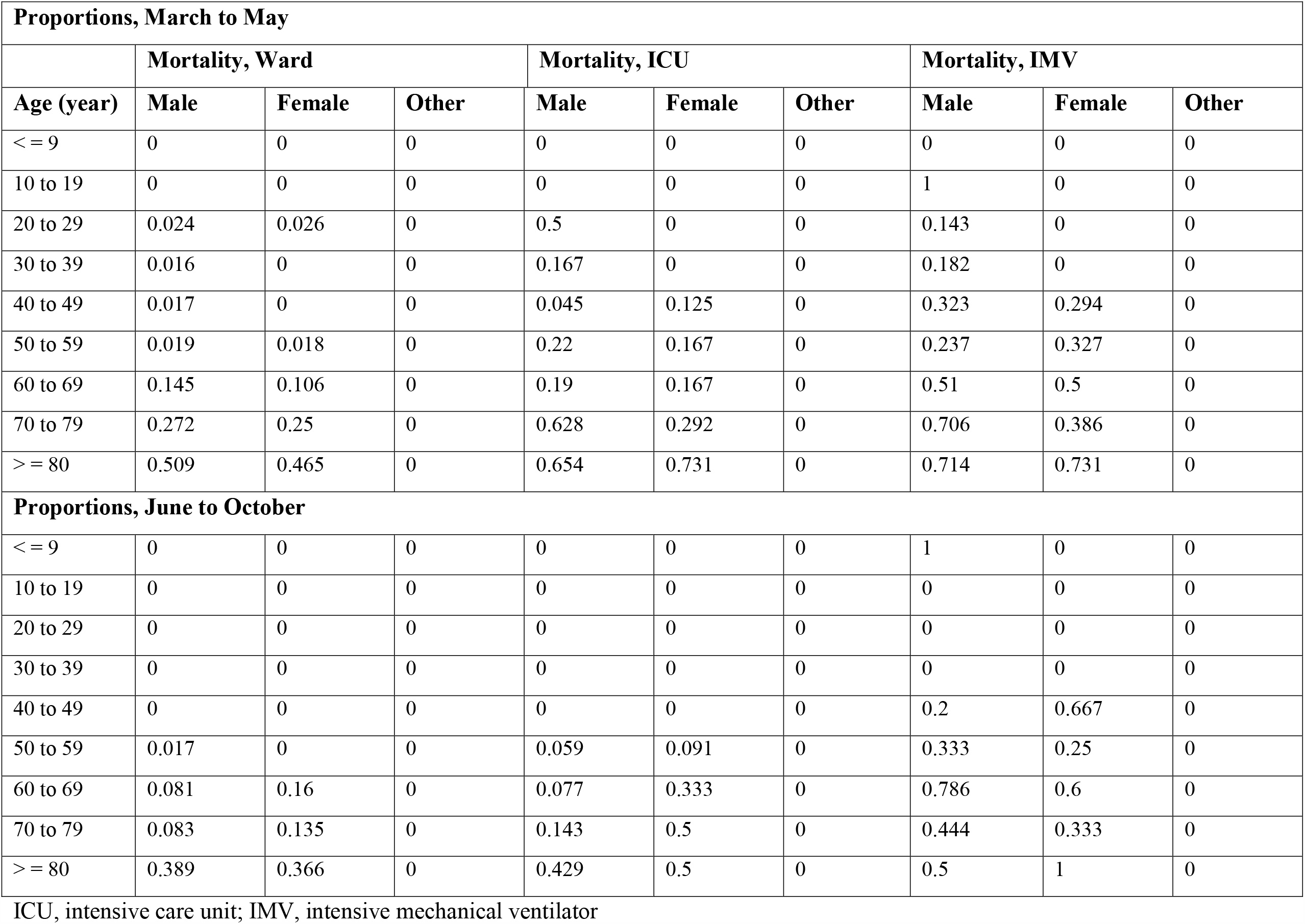
Mortality by acute care level, age and sex.

### Length of Stay

We updated LOS (days) for each acute care level, which also changed over time by month **(Table 4)**. When the individual is hospitalized, LOS is sampled from poisson and gamma distributions informed by monthly mean LOS and standard deviation. For patients who are admitted to ward, the average LOS was between 5.7 and 16.15 days. For patients requiring ICU care without ventilation, their average stay in the ICU was between 6.5 to 10.7 days. For patients who needed ICU care and ventilation, their average stay in the ICU was between 14.8 and 21.6 days. For any individual requiring ICU care (ventilator or no ventilator), the time spent in the ward prior and post ICU admission was 6.83 days; we assumed the 6.83 days was equally divided for the ward LOS pre- and post-ICU. Individuals requiring ventilators spend on average 1.2 days in the ICU without ventilation. Overall, for an individual requiring ventilation, their LOS includes the following distinct components: time spent in the ward prior and post to ICU admission, time in the ICU not ventilated, plus time on ventilation. Anytime during this LOS, patients may die. When an individual no longer requires a resource, either due to completion of LOS and discharge or death, the resource will become available to other individuals. Individuals who cannot access their required resource form a queue and may die depending on their disease severity.

**Table 4.**
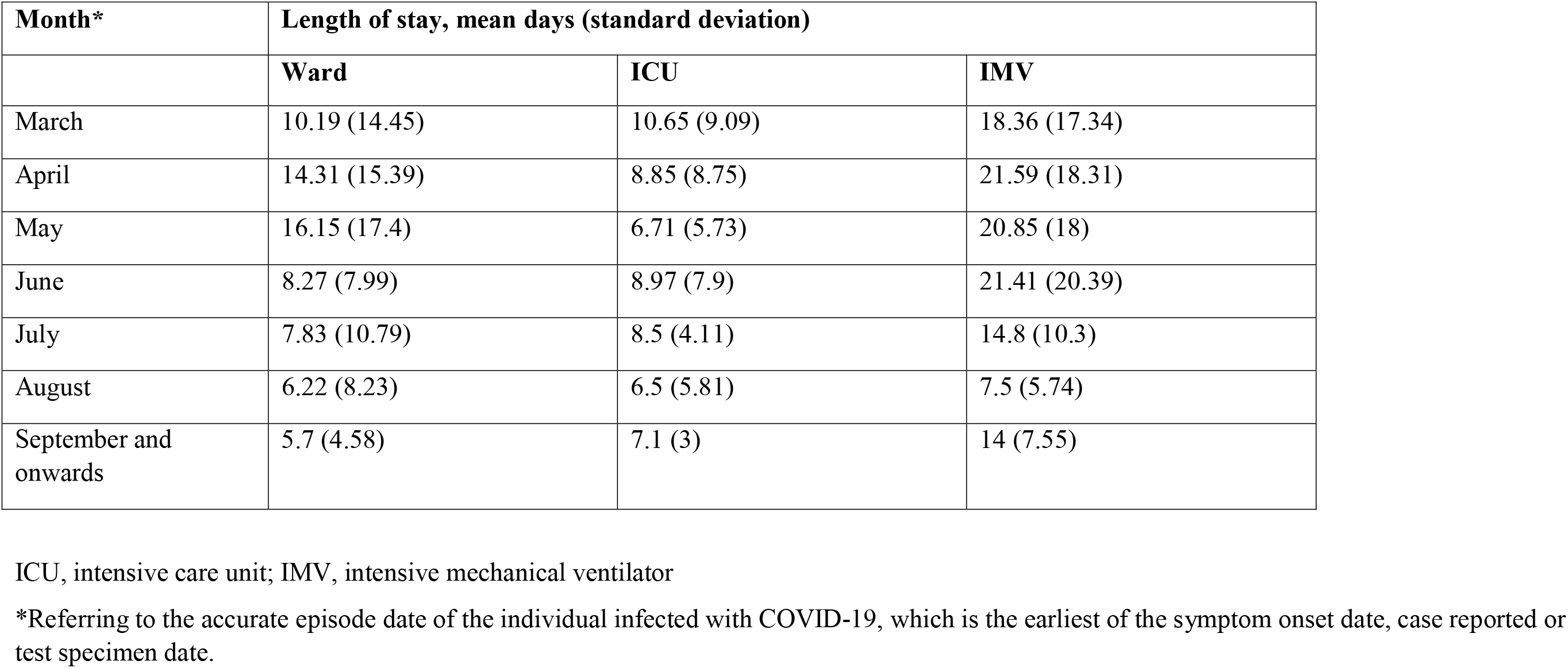
Length of stay over time by acute care level.

| Month* | Length of stay, mean days (standard deviation) |  |  |
| --- | --- | --- | --- |
|  | Ward | ICU | IMV |
| March | 10.19 (14.45) | 10.65 (9.09) | 18.36 (17.34) |
| April | 14.31 (15.39) | 8.85 (8.75) | 21.59 (18.31) |
| May | 16.15 (17.4) | 6.71 (5.73) | 20.85 (18) |
| June | 8.27 (7.99) | 8.97 (7.9) | 21.41 (20.39) |
| July | 7.83 (10.79) | 8.5 (4.11) | 14.8 (10.3) |
| August | 6.22 (8.23) | 6.5 (5.81) | 7.5 (5.74) |
| September and onwards | 5.7 (4.58) | 7.1 (3) | 14 (7.55) |

ICU, intensive care unit; IMV, intensive mechanical ventilator
\*Referring to the accurate episode date of the individual infected with COVID-19, which is the earliest of the symptom onset date, case reported or test specimen date.

### Analysis and Calibration

The model starts simulations on March 1, 2020. The number of COVID-19 cases between March 1, 2020 and October 31, 2020 were taken from Ontario’s Ministry of Health data.(6) We calibrated some LOS components (ICU, non-ventilated) to reported ward and ICU (with and without ventilation) occupancy between June 15 and October 31, 2020.(7) The non-ventilated ICU LOS was calibrated only for the most recent month of October since the analysis is subject to some censoring (i.e., individuals who may not have an outcome at the end of analysis follow-up time). The simulated occupancy from CORE 2.0 is compared to the reported occupancy data for all levels of care in **Figures 2a-2c**. Overall, simulations for all three level of care resources compare well with reported occupancy data up until October 31, 2020. A brief summary of all parameters is in **Table 5**.

**Table 5.** Summary of model inputs for CORE 2.0.

| **Acute Care Resource Use** | **Probability** | **Sources** |
| --- | --- | --- |
| Ward | 0 to 0.35<br>[varies with age and sex] | Mac 2020 (4)<br>[Calibration estimate] |
| ICU | 0 to 0.38<br>[varies with age, sex, and month] |  |
| Ventilation | 0 to 0.73<br>[varies with age and sex] |  |
| **Length of Stay** | **Days (mean)** |  |
| Ward | 5.7 to 16.15<br>[varies by month] | Mac 2020 (4)<br>[Calibration estimate] |
| Ward (pre-ICU or post-ICU) | 6.83 |  |
| ICU (Non-vent) | 6.5 to 10.65<br>[varies by month] |  |
| ICU (Pre-vent) | 1.20 |  |
| ICU (Vent) | 7.5 to 21.59<br>[varies by month] |  |
| **Mortality** | **Rate** |  |
| Ward | 0 to 0.51<br>[varies with age, sex, and month] | Mac 2020 (4) |
| ICU (Non-vent) | 0 to 0.73<br>[varies with age, sex, and month] |  |
| ICU (Vent) | 0 to 0.79<br>[varies with age, sex, and month] |  |

**Figure 2a.**
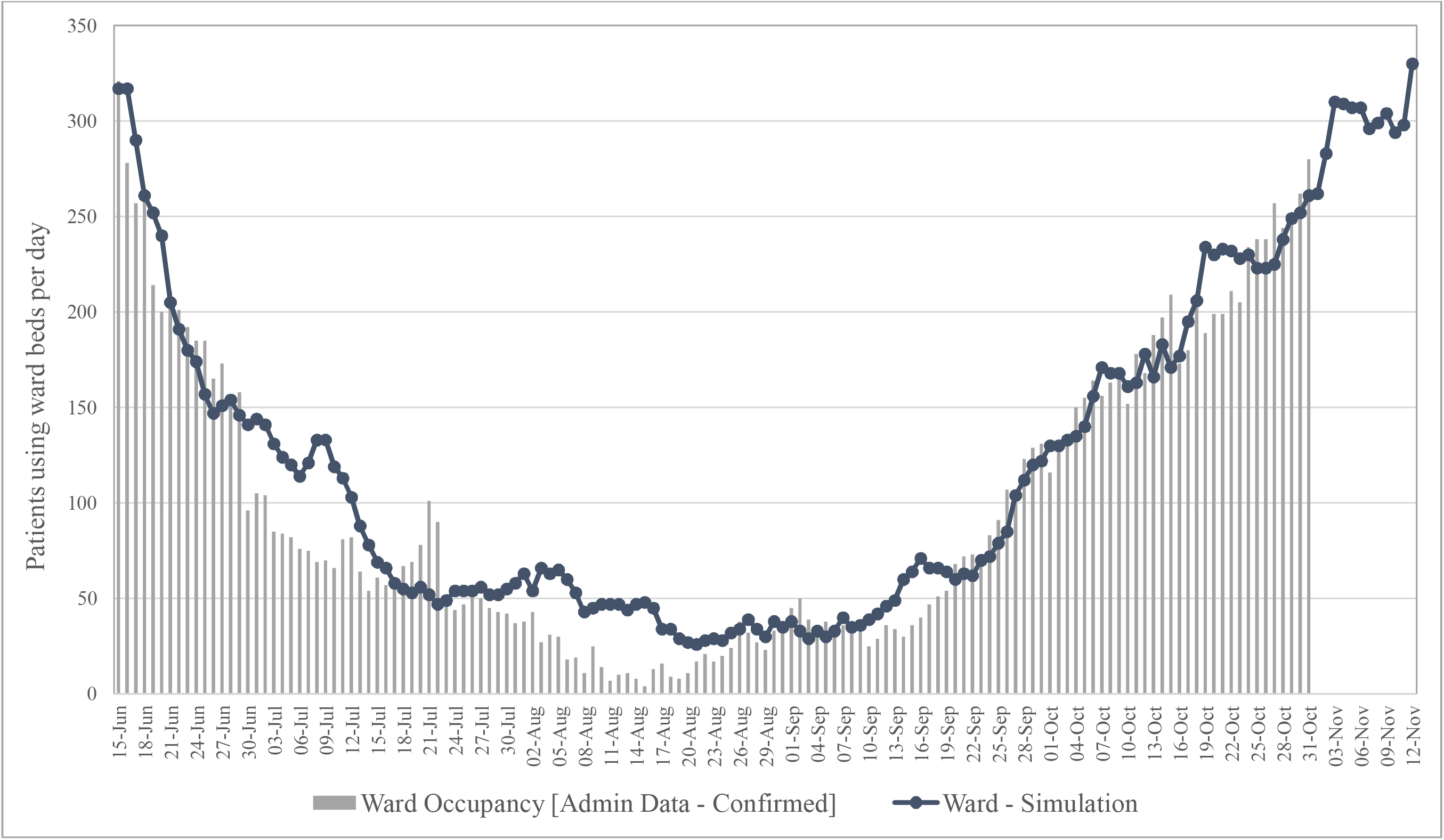
Predicted and confirmed ward occupancy in Ontario based on administrative data.

**Figure 2b.**
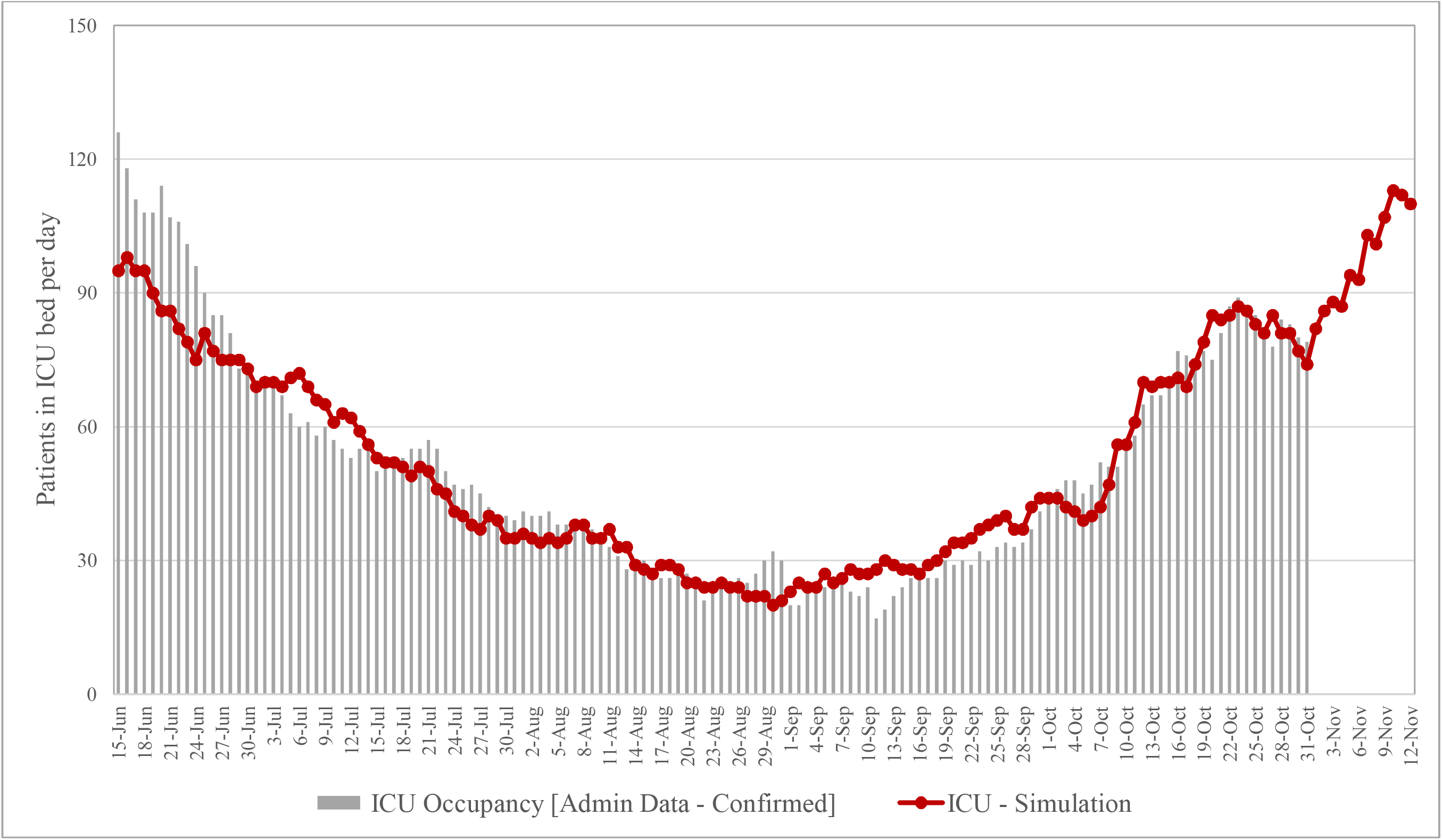
Predicted and confirmed ICU occupancy in Ontario based on administrative data.

**Figure 2c.**
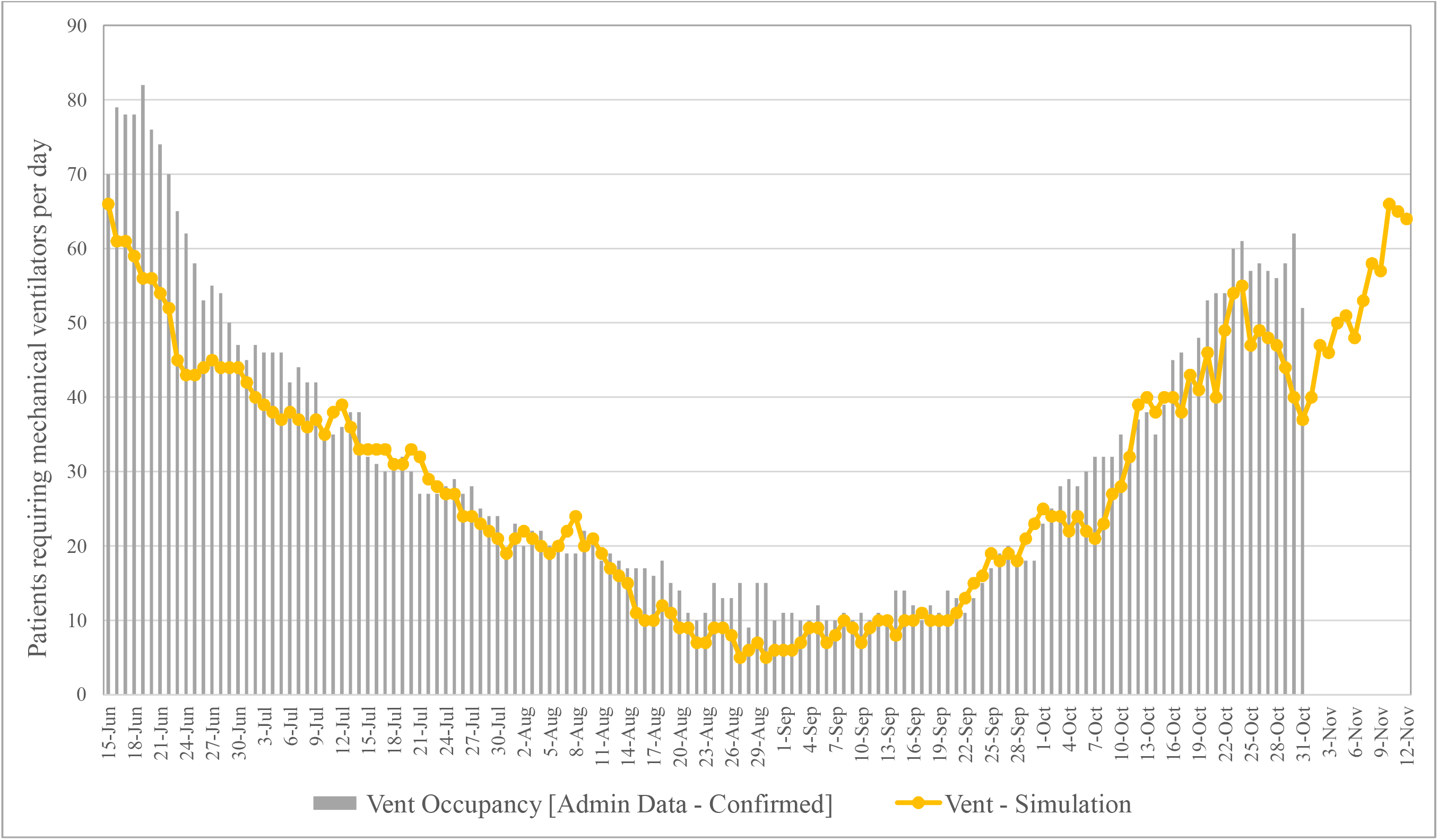
Predicted and confirmed ventilator occupancy in Ontario based on administrative data.

**Figure 3.**
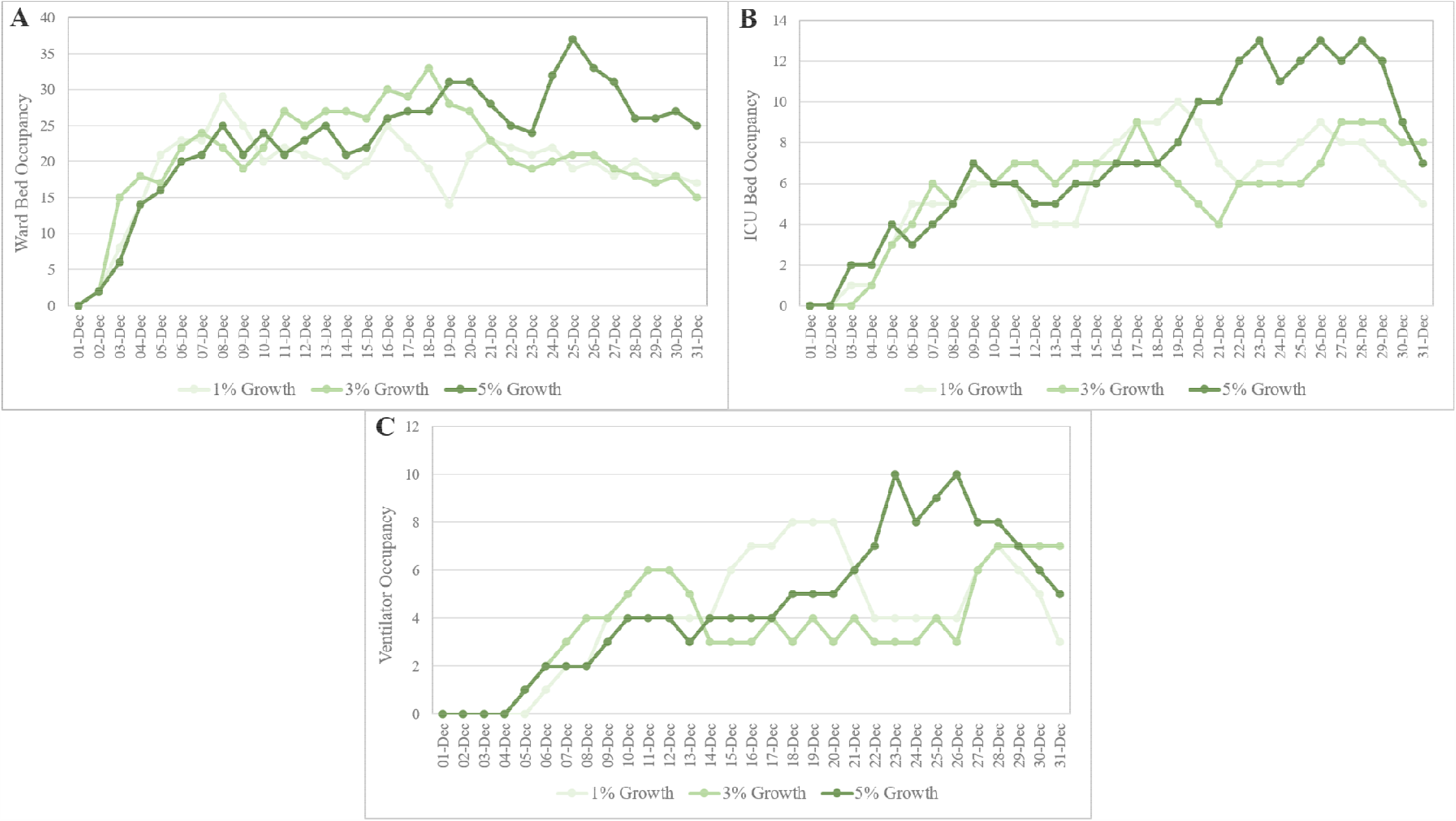
Predicted resource use in Waterloo region between Dec 1 and 30, 2020 for A) Ward beds B) ICU beds C) Ventilators.

### Regional Application of CORE 2.0 in Ontario

Here we describe an example of the use of CORE 2.0 for a regional analysis of acute care resource use for the Region of Waterloo, Ontario, Canada that serves 617,870 people from the cities of Cambridge, Kitchener, Waterloo and townships of North Dumfries, Wellesley, Wilmot and Woolwich. In this example, we simulate the ward bed, ICU bed, and ventilator occupancies for 30 days starting on December 1, 2020 for three different case trajectory scenarios:

1. Growth 1.01: Growth of incident cases for 10 days at 1%, followed by contraction (reduction) of incident cases by 5% (i.e., [incident cases] _day_ = [incident cases] _day-1_*0.95, where day = current date) for the remainder of the 30 days.
2. Growth 1.03: Growth of incident cases for 10 days at 3%, followed by contraction (reduction) of incident cases by 5% for the remainder of the 30 days.
3. Growth 1.05: growth of incident cases for 10 days at 5%, followed by contraction (reduction) of incident cases by 5% for the remainder of the 30 days.

These scenarios were chosen based on input from stakeholders from the region, and are designed to capture the potential decrease in cases after recent implementation of public health interventions to limit community transmission. The contraction rate of 5% starting at 10 days after peak are user-defined assumptions. The model used a run-in time of 8-days prior to the simulation start date (i.e., November 23, 2020), allowing the reported cases in the past 8 days to also start being hospitalized (if needed) on December 1^st^, and onwards, to provide a more accurate number of occupancies required within the next 30 days. The reported cases for November 23 to 30, 2020 were obtained from the Region of Waterloo.(8) On November 30, 2020, there were 66 incident cases reported, which is what the case growth scenarios started with; all reported cases used as input is reported in **Table 6**.

**Table 6.** Case inputs for regional example.

|  | Contraction | Growth |  |  |
| --- | --- | --- | --- | --- |
|  | 0.95 | 1.01 | 1.03 | 1.05 |
| Date |  | Scenario 1 | Scenario 2 | Scenario 3 |
| 23-Nov | As reported by Waterloo region | 62 | 62 | 62 |
| 24-Nov |  | 78 | 78 | 78 |
| 25-Nov |  | 67 | 67 | 67 |
| 26-Nov |  | 74 | 74 | 74 |
| 27-Nov |  | 90 | 90 | 90 |
| 28-Nov |  | 77 | 77 | 77 |
| 29-Nov |  | 57 | 57 | 57 |
| 30-Nov |  | 66 | 66 | 66 |
| 01-Dec | Growth as indicated in the scenario (1%, 3%, or 5%) | 66 | 66 | 66 |
| 02-Dec |  | 67 | 68 | 69 |
| 03-Dec |  | 67 | 70 | 73 |
| 04-Dec |  | 68 | 72 | 76 |
| 05-Dec |  | 69 | 74 | 80 |
| 06-Dec |  | 69 | 77 | 84 |
| 07-Dec |  | 70 | 79 | 88 |
| 08-Dec |  | 71 | 81 | 93 |
| 09-Dec |  | 71 | 84 | 98 |
| 10-Dec |  | 72 | 86 | 102 |
| 11-Dec | Contraction (5%) | 69 | 82 | 97 |
| 12-Dec |  | 65 | 78 | 92 |
| 13-Dec |  | 62 | 74 | 88 |
| 14-Dec |  | 59 | 70 | 83 |
| 15-Dec |  | 56 | 67 | 79 |
| 16-Dec |  | 53 | 63 | 75 |
| 17-Dec |  | 50 | 60 | 72 |
| 18-Dec |  | 48 | 57 | 68 |
| 19-Dec |  | 45 | 54 | 65 |
| 20-Dec |  | 43 | 52 | 61 |
| 21-Dec |  | 41 | 49 | 58 |
| 22-Dec |  | 39 | 47 | 55 |
| 23-Dec |  | 37 | 44 | 53 |
| 24-Dec |  | 35 | 42 | 50 |
| 25-Dec |  | 33 | 40 | 47 |
| 26-Dec |  | 32 | 38 | 45 |
| 27-Dec |  | 30 | 36 | 43 |
| 28-Dec |  | 29 | 34 | 41 |
| 29-Dec |  | 27 | 32 | 39 |
| 30-Dec |  | 26 | 31 | 37 |
| 31-Dec |  | 25 | 29 | 35 |

The results reported are the ward bed, ICU bed, and ventilator occupancies in **Figures 3A-3C**. These are the number of people requiring this resource on specific dates, and not the new admissions (i.e., there are new admissions, and also discharges based on survival, and length of stay). In these scenarios, as the increase of incident cases grows faster, the need for all resources increases. In the most conservative scenario used to estimate the capacity required, the Region of Waterloo may require up to 29 ward beds, 10 ICU beds, and 8 ventilators for COVID-19 patients, and in the worst case growth estimate (5%), these numbers increase to 37, 13, and 10, respectively.

There are some limitations to this regional analysis. First, the model parameters have all been calibrated and validated to Ontario’s population-level use of COVID-19 resource use, length of stay, and mortality by age and sex. This does not necessarily reflect the population from the Region of Waterloo. However, the Region of Waterloo is not considered a rural area or geographically different from many of the hotspots in Ontario (e.g., Toronto, Peel Region) and so the demographics of COVID-19 cases should still apply. Second, the results represent the occupancy that would be seen in the entire Region of Waterloo region, and not necessarily at a specific hospital. These results do not consider the current ward, ICU and ventilator occupancies prior to December 1, 2020. Unfortunately, because these individuals’ trajectory in their LOS is unknown, it was not possible to add these current occupancies into the model and so these must be considered for the near-term future in addition to the new resources predicted. Despite these caveats, CORE 2.0 was able to simulate acute care resource needs for regional analysis and provide an estimate for local hospital planners for resource allocation purposes.

## Conclusion

CORE 2.0 now simulates individuals and their COVID-19 disease trajectory incorporating age, sex, and temporal trends for acute care resource use, mortality, and LOS. This model is highly flexible and customizable to data updates, and can better inform acute care planning and public measures as the pandemic progresses.

## Data Availability

All data is presented in the Tables and Appendix of the manuscript and referenced when cited from the literature.

## Appendix

### Appendix 1. Patient Pathways

1. Sent home: We assume individual self-isolates and recovers; these patients are not explicitly modeled.
2. Admitted to ward: The individual is assigned a LOS sampled from a distribution that determines their LOS in the hospital. This individual is now subject to a daily probability of death. Once their stay is complete (i.e., did not die), this individual is discharged home and recovers.
3. Admitted to intensive care unit (ICU): This individual is assigned a LOS sampled from a distribution for their ward stay (pre-ICU). If this sampled LOS for the ward (pre-ICU) stay is < 1, this individual proceeds straight to the ICU on the same day. If the LOS for the pre-ICU ward stay ≥ 1, then the individual is first admitted to the ward and they will deteriorate at a later day to require ICU admission.
  - This new structure more accurately simulates patient pathway, so that they can enter straight to ICU, or deteriorate later.
  - Once this individuals’ stay in the ward is equal to the LOS sampled for their ward stay (pre-ICU), this individual would deteriorate and are admitted to the ICU if resources are available. This individual is then assigned a LOS, sampled from a distribution, for their ICU admission and have a daily probability of death in the ICU.
  - Once their stay is complete (i.e., did not die), they are assigned a LOS in the ward (post-ICU), sampled from a distribution, and spend their recovery in the ward.
4. Admitted to ICU with mechanical ventilator: If an individual is admitted to the ICU, there is a probability they will require a ventilator. If they do not need a ventilator, then they continue to the ICU like Step 3 above.
  - If an individual requires a ventilator, they are assigned a LOS, sampled from a distribution, to determine the number of days they stay in the ICU before requiring ventilation (pre-Vent). If this LOS < 1, this individual will require a ventilator immediately and receive it if the resources are available.
  - If this pre-Vent LOS ≥ 1, then this individual is admitted to the ICU and does not require a ventilator yet. Once this individuals’ stay in the ICU is equal to the pre-Vent LOS, they will deteriorate and require a ventilator.
  - This new structure more accurately simulates patient pathway, so that they can need a ventilator right away, or deteriorate while in the ICU later.
  - If a ventilator is available, they will be assigned a LOS sampled from a distribution. If they recover, they will move to the ward if these resources are available.

## References

1. Barrett K, Khan YA, Mac S, Ximenes R, Naimark DMJ, Sander B. Estimation of COVID-19– induced depletion of hospital resources in Ontario, Canada. Can Med Assoc J. 2020 May 14;192(24):E640–646.

2. Mac S, Ximenes R, Barrett K, Khan YA, Naimark D, Sander B. Predicting Hospital Resource Use in Ontario For a Potential Second Wave [Internet]. 2020. Available from: www.covid-19-mc.ca

3. Public Health Ontario. iPHIS Resources [Internet]. 2020 [cited 2020 Oct 9]. Available from: https://www.publichealthontario.ca/en/diseases-and-conditions/infectious-diseases/ccm/iphis

4. Mac S, Barrett K, Khan YA, Naimark DMJ, Rosella L, Ximenes R, et al. COVID-19 Demographics, Acute Care Resource Use and Mortality by Age and Sex in Ontario, Canada: Population-based Retrospective Cohort Analysis. medRxiv [Internet]. 2020 Jan 1;2020.11.04.20225474. Available from: http://medrxiv.org/content/early/2020/11/06/2020.11.04.20225474.abstract

5. Mac S, Barrett K, Khan YA, Naimark DMJ, Rosella L, Ximenes R, et al. COVID-19 Demographics, Acute Care Resource Use and Mortality by Age and Sex in Ontario, Canada: Population-based Retrospective Cohort Analysis. medRxiv. 2020 Jan;2020.11.04.20225474.

6. Government of Ontario. COVID-19 case data: All Ontario [Internet]. 2020 [cited 2020 Oct 9]. Available from: https://covid-19.ontario.ca/data

7. Ontario Ministry of Health. Daily Bed Census. 2020.

8. Region of Waterloo. Waterloo Region COVID-19 summary [Internet]. 2020 [cited 2020 Dec 9]. Available from: https://www.regionofwaterloo.ca/en/health-and-wellness/positive-cases-in-waterloo-region.aspx#

